# Ohio Amish may be disproportionately at risk for La Crosse virus infection

**DOI:** 10.64898/2026.01.14.26344083

**Authors:** Morgan E. Chaney, Christina M. Bergey

## Abstract

La Crosse virus (LACV) causes the majority of pediatric arboviral encephalitis cases in the United States, with Ohio historically reporting the highest incidence of LACV neuroinvasive disease (LACV-ND). To identify county-level risk factors, we analyzed two decades (2003-2023) of Ohio surveillance data across all 88 counties using spatial statistical methods. Land-use correlation screening identified deciduous forest cover (ρ = 0.36, adjusted p = 0.006) and pasture/hay cover (ρ = 0.37, adjusted p = 0.004) as significant environmental correlates. However, Amish population share emerged as a substantially stronger predictor (ρ = 0.71, p << 0.001). Cases clustered in east-central Ohio, where Holmes County, home to the world’s second-largest Amish settlement, exhibited the highest statewide incidence. We fit a Spatial Durbin Model to decompose within-county (direct) and spillover (indirect) effects while controlling for spatial autocorrelation. Amish population share remained the sole significant predictor after adjusting for land use and spatial structure, with total effects (β = 8.2) significantly larger than environmental variables (deciduous forest β = 0.3, pasture/hay β = -0.7). These findings indicate that Amish lifeways, potentially including occupational exposures, housing characteristics, or healthcare-seeking behaviors, represent critical but understudied LACV risk factors. Given the severe morbidity of pediatric LACV-ND and absence of vaccines, our results support targeted prevention efforts in Ohio Amish communities, contingent on culturally appropriate intervention strategies.

## Introduction

La Crosse virus (LACV) is a mosquito-borne orthobunyavirus and a leading cause of pediatric arboviral encephalitis in the United States. A subset of infections progress to LACV neuroinvasive disease (LACV-ND), characterized by encephalitis and associated symptoms like seizures ^1–5^, with death occurring in 1-2% of cases ^2,6^. LACV infections have been increasing in central and southern Appalachia ^7^, with hotspots in West Virginia, Tennessee/North Carolina, Wisconsin, and Ohio ^8^. Given the lack of a vaccine, reducing pediatric disability and deaths due to LACV requires identifying and disrupting the fine-scale ecological and behavioral drivers of transmission.

LACV transmission is maintained primarily in hardwood forests by the native eastern treehole mosquito, *Aedes triseriatus* ^1,2^, with documented accessory transmission by congeners [e.g., *Ae. albopictus*: ^9^; *Ae. japonicus*: ^10^]. Female mosquitoes acquire infection chiefly when blood-feeding on viremic amplifying hosts, notably chipmunks (*Tamias striatus*) and tree squirrels (*Sciurus spp.*), although *Ae. triseriatus* mosquitoes can also be infected venereally or vertically (reviewed by ^2,7,11^). Large-bodied mammals including humans are tangential or dead-end hosts due to their lower levels of viremia ^1,2,12^. Most human infections are mild or asymptomatic ^1^, and therefore severely under-reported. Detection likely only typically occurs in the small proportion of infections that develop into LACV-ND. LACV-ND cases occur typically in young children (4-11 years old) ^1,2^, when their state of active neurogenesis may increase susceptibility to LACV-induced apoptosis (Winkler et al. 2019) and the integrity of the blood-brain barrier is naturally lower ^13^.

The ecological determinants of LACV spillover to humans remain incompletely characterized. *Ae. triseriatus* mosquitoes reproduce in tree holes or similar artificial containers, and the presence of maple trees and artificial containers has been positively associated with these mosquitoes ^14^. Forest disturbance has been found to negatively correlate with LACV prevalence, with less-disturbed sites harboring the highest abundance of vectors *Ae. triseriatus* ^15,16^ and *Ae. japonicus* ^15^ and highest rates of LACV-infected mosquitoes ^17^. Similarly, a study of pairs of adjacent forested and peridomestic habitats found *Ae. triseriatus* to be most prevalent in the forests ^18^. However, the habitat preference was inverted in locations characterized by a high density of artificial containers (“poorly maintained peridomestic habitats”), with the mosquitoes instead most commonly found near the houses. This finding suggests that built-environment features might restructure breeding habitats and pull vector populations into peridomestic settings where contact with children is more likely, thereby increasing spillover risk.

Vector presence alone does not explain LACV occurrence, with LACV-ND distribution highly focal despite the near ubiquity of competent LACV vectors across most of the eastern United States ^7^. Work from endemic regions points to a multilevel set of drivers, including precipitation and temperature ^19^, vapor pressure deficit index, proportion of children, and land-use categories ^20^.

Serologic studies of people living in endemic areas often indicate surprisingly high rates of prior LACV infection ^21–29^. For example, people living on the Cherokee Indian Reservation of western North Carolina had a LACV seropositivity rate of 20.6%. This high rate contrasted with lower seropositivity in nearby locations outside of the reservation [2.5% – 8.3%; ^27^]. Such results [see also ^30^] suggest that micro-geographic community-level contexts, perhaps including differences in built environment or human behavior, may concentrate LACV exposure in specific populations living alongside lower-risk neighbors. However, the drivers of this heterogeneity remain poorly characterized.

Ohio has the highest number of reported LACV-ND cases since 2003 ^31^, and against this backdrop, Holmes County is an extreme outlier. Since 2003, Holmes has had an average LACV-ND incidence of 2.95 cases per 100,000 people, several-fold higher than all other Ohio counties ^31^. This pattern raises a central question: Is the concentration of LACV-ND cases in Holmes County simply the expected tail of current habitat-suitability models (forest cover, tree-hole resources, temperature/precipitation, housing density), or does Holmes represent an exception to those rules? If the latter, a plausible hypothesis is that population structure and culturally patterned behaviors concentrated in Holmes modulate exposure and disease.

Holmes has the highest concentration of Old Order Amish of any U.S. county (approximately 45.5% of total county residents in 2020), offering a natural experiment to examine how household infrastructure, outdoor and agricultural activities, and preventive practices might alter mosquito contact and infection rates.

Here, we analyze Ohio surveillance and environmental data to: i.) quantify how well standard habitat variables account for county-level variation in LACV-ND incidence, ii.) test whether Holmes County’s burden is predicted by those variables or remains an outlier, and iii.) explore whether community-level factors (including the proportion of Old Order Amish residents as a proxy for distinct household and occupational patterns) improve model fit.

## Materials and Methods

We downloaded LACV-ND average annual incidence per 100,000 residents by county of residence spanning the period between 2003 and 2024 ^31^. Because this variable’s distribution showed a pronounced rightward skew, we then log-transformed our LACV-ND incidence rate.

For estimates of the Ohio Amish population, we used data from the 2020 U.S. Religion Census ^32^. In filtering these data, we included only groups with the keyword *Amish* in their denomination name (e.g., Beachy Amish-Mennonite, Berea Amish-Mennonite), but the largest category for the Ohio Amish was simply the undifferentiated Amish denomination, comprising 97.2% of the population (i.e., 71,756 out of 73,792 adherents).

For our data on land use, we used the most recent version of the National Land Cover Database [NLCD; ^33^]. This raster was downloaded through ArcGIS Pro, and we used the *Model Builder* tool to automate (1) the clipping of this raster based on generalized US counties shapefile and (2) the tabulation of each Ohio county’s proportional area based on the NLCD land-use categories (**Table 1**). All of the resulting tables were joined into a combined spreadsheet outside of ArcGIS prior to being merged with all of the abovementioned demographic data based on each county’s Federal Information Processing Standard (FIPS) code. All demographic data were merged with the ArcGIS-derived land-use data.

**Table 1:**
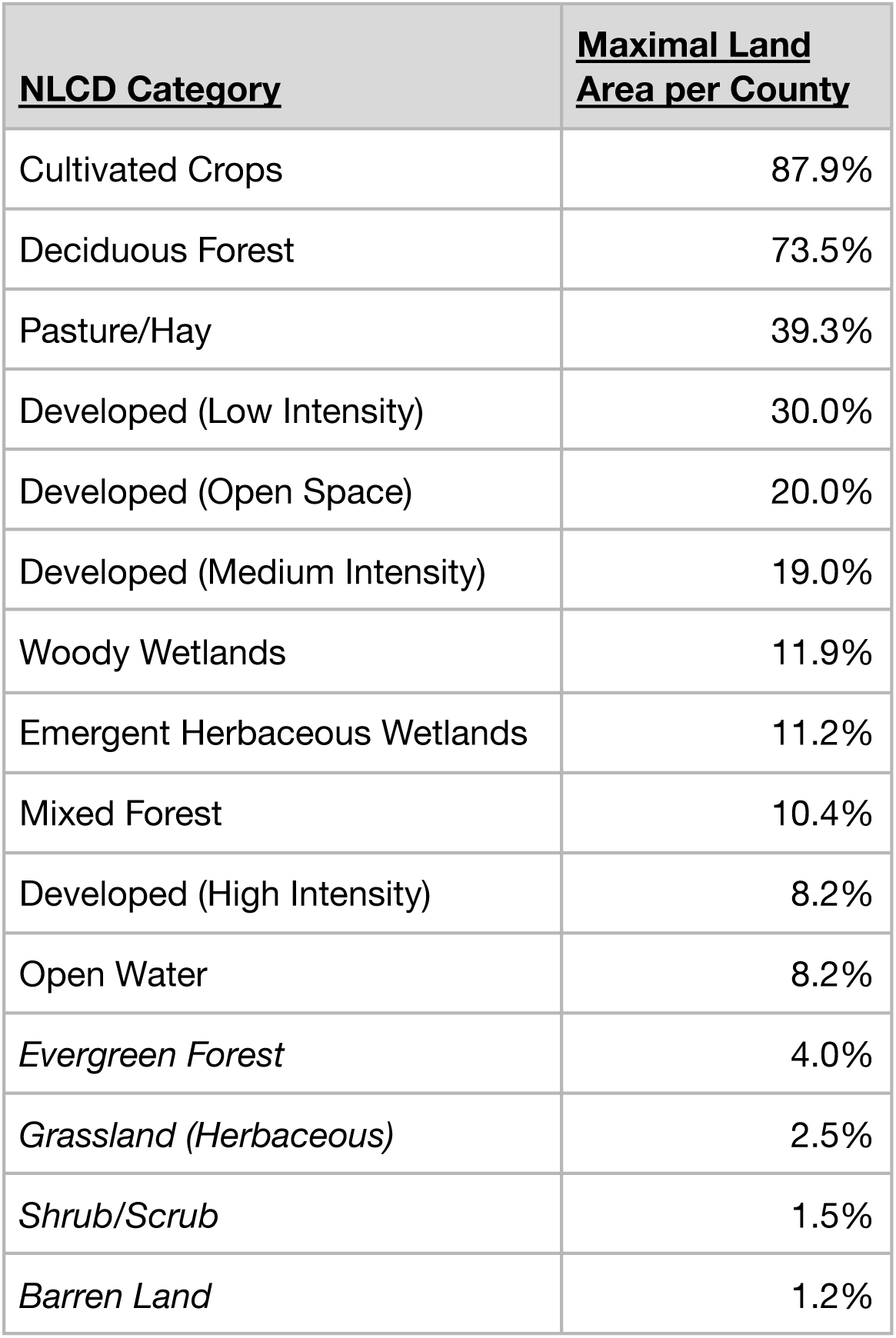
Variables measured by the National Land Cover Database (Dewitz, 2019) and used in this analysis. We did not consider the italicized categories for any further modeling because of their absolutely low representation in the dataset (<5% maximal land area per county).

As a starting point, we performed a correlation screen using Spearman’s ρ of all land-use categories in the NLCD against the LACV-ND case rate. Three land-use types showed a significant correlation (i.e., barren land, evergreen forest, and shrublands), but we disregarded these land-use categories downstream because none exceeded 5% of the land area in any county. Due to the geographic nature of these data, and because our response LACV-ND rate showed significant spatial autocorrelation (Moran’s *I* = 0.35, *p* =1.42e-9), we opted to use a spatial modeling strategy. We used the significant correlates from our abovementioned screen for constructing these spatial models using the *spatialreg* R package ^34^. We evaluated which type of spatial model would be most appropriate using Rao’s score from the *spdep* R package ^35,36^. In all cases, these tests indicated significant results for both error and lag models; therefore, we opted to use spatial Durbin models for this analysis.

Spatial Durbin models account for spatial autocorrelation by including lagged variables for each land-use category based on its neighboring counties [reviewed by ^37^]. To do this, we generated a weighting matrix (*W*) that described all of the direct neighbors for each Ohio county with weights equal to 1.00 / *C*, where *C* is the number of counties sharing a border with the county in question. This matrix was used to calculate each county’s lagged value (WX)*_i_* for each variable as:

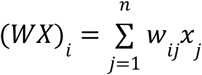

where *w_ij_* is the weighted value mentioned above and *x_j_* is the direct value of the variable in question for each neighboring county. In essence, this calculation provides an average of all of a county’s neighbors while excluding the direct value of the county itself, which is already included in the model by the direct value of each county.

After incorporating each county’s proportional Amish population, we constructed ordinary least-squares linear models primarily to check for multicollinearity of these predictor variables. The variance inflation factors (VIFs) for these terms were all acceptable (range: 1.06, 2.12), and we therefore concluded no substantive amount of multicollinearity existed in our subsequent models.

## Results

Across Ohio’s 88 counties, LACV-ND incidence varied substantially, ranging from 0 to 2.95 cases per 100,000 population (**Fig. S1**). The presence of Amish populations was strongly associated with LACV-ND occurrence: all 42 counties with non-zero Amish populations recorded at least one LACV-ND case during the study period, whereas only 25 of 46 counties (54%) without Amish populations reported any cases (**Fig. S2**). Both Amish population share and LACV-ND incidence exhibited clear geographic clustering in east-central and northeastern Ohio, with Holmes County representing an extreme outlier at 46% Amish population share and the highest LACV-ND incidence statewide (**Fig. S1, S3**).

To investigate how LACV-ND incidence covaried with environmental variables, we performed a univariate screen by computing the Spearman correlation coefficient for each of the land-use categories tracked within Ohio counties in the NLCD. Because three categories (i.e., barren land, evergreen forest, and shrublands) accounted for less than 5% of land area for every county, we did not consider results from them in downstream analysis. Of the remaining 11 land-use variables, two environmental variables correlated with LACV-ND incidence (**Table 2**): proportional land area devoted to pasture/hay (ρ = 0.37, *p_adj_* = 0.004) and deciduous forests (ρ = 0.36, *p_adj_* = 0.006).

**Table 2:** Summary of correlations performed with Spearman’s ρ (non-directional hypothesis testing for all). Raw *p*-values are displayed; we used a factor of 11 when correcting these for multiple comparisons. Bolded rows represent categories that we used for subsequent spatial Durbin models.

| **NLCD Land-use Category** | **Spearman's $\rho$** | **Raw $p$-value** |
| --- | --- | --- |
| Open Water | -0.19 | 0.07 |
| Developed (Open Space) | 0.03 | 0.81 |
| Developed (Low Intensity) | -0.26 | 0.01 |
| Developed (Medium Intensity) | -0.22 | 0.04 |
| Developed (High Intensity) | -0.24 | 0.02 |
| **Deciduous Forest** | **0.35** | **0.0009** |
| Mixed Forest | 0.21 | 0.05 |
| **Pasture/Hay** | **0.36** | **0.0005** |
| Cultivated Crops | -0.21 | 0.05 |
| Woody Wetlands | 0.22 | 0.13 |
| Emergent Herbaceous Wetlands | -0.03 | 0.76 |

Next, we investigated whether including the proportion of Old Order Amish residents as a proxy for distinct household and occupational patterns improves model fit. We found a strong positive correlation between LACV-ND cases and the countywide proportional Amish population (Spearman’s ρ = 0.71, *p* << 0.001), even after excluding Holmes County from the dataset (ρ = 0.70, *p* << 0.001; (**Fig. 1**). This relationship remained robust whether or not Holmes County was included in the fit, suggesting the association is not driven solely by this single high-leverage observation. The relationship between Amish share and LACV-ND incidence was evident across the full range of Amish population densities: counties in the highest quartile of non-zero Amish share (Q4) showed substantially elevated incidence compared to counties with zero Amish populations, with Holmes County representing a clear outlier within Q4 (**Fig. 2**).

**Figure 1:**
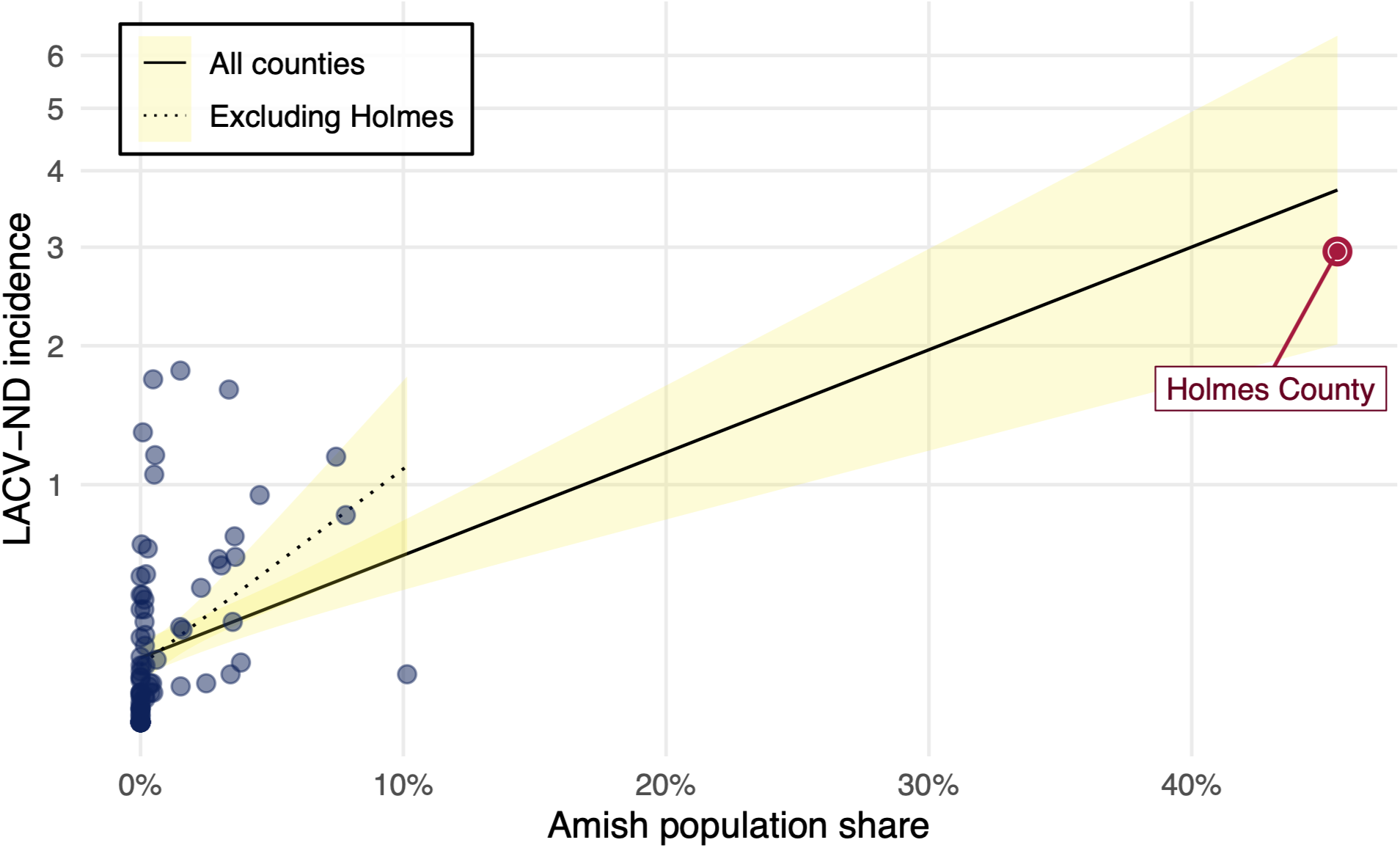
Association between Amish population share and LACV-ND incidence. Scatterplot of county-level data with Amish share (x-axis) versus LACV-ND incidence (y-axis, log_1+_-transformed but displayed on original scale). Two ordinary least squares regression lines are overlaid: solid line includes all counties; dashed line excludes Holmes County. Holmes County is highlighted as a distinct point annotation.

**Figure 2:**
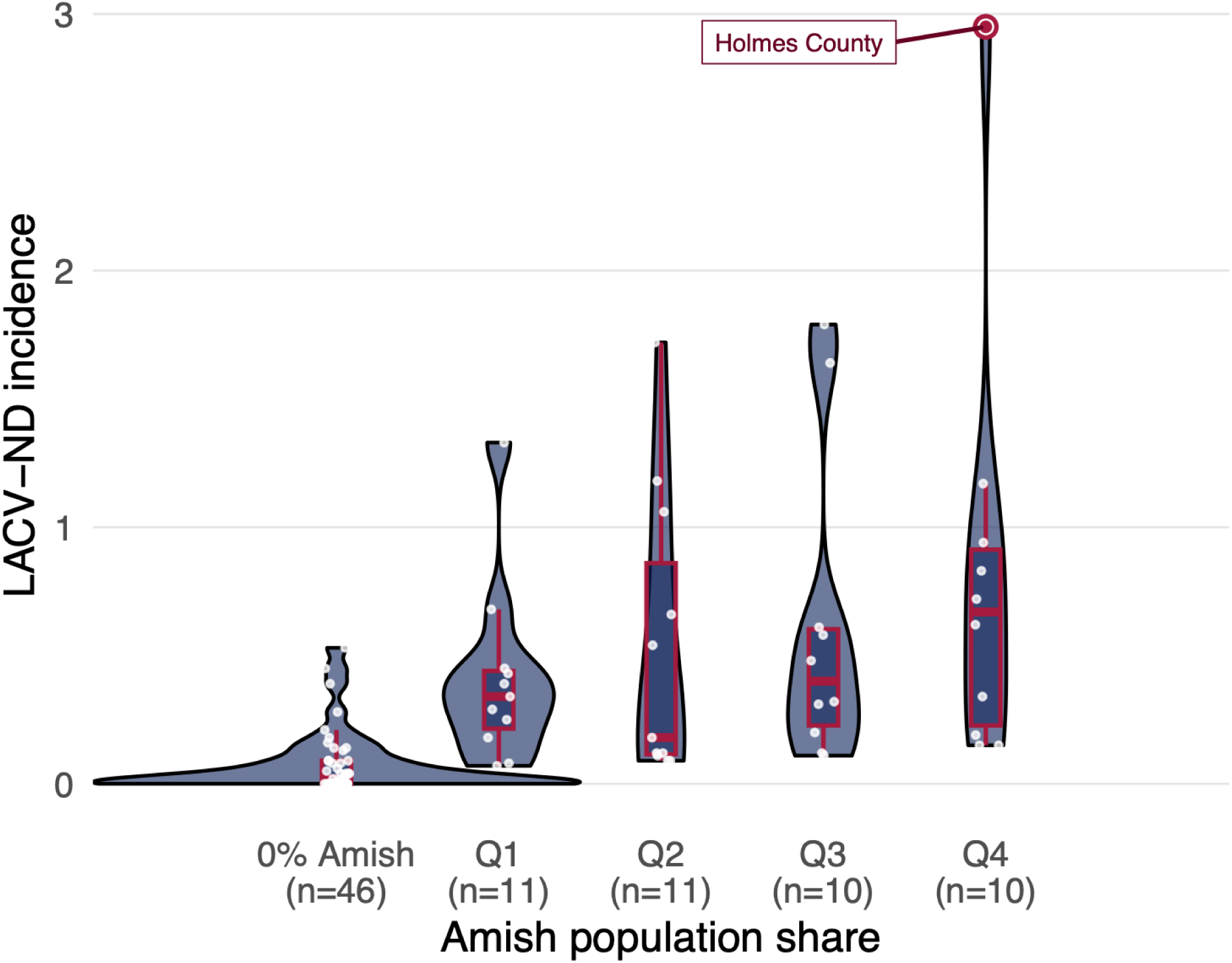
LACV-ND incidence by Amish population share. Violin plots show the kernel density distribution of county-level incidence, with overlaid boxplots (median and interquartile range) and individual counties shown as semi-transparent points. Counties are grouped into five categories: 0% Amish share, and quartiles Q1-Q4 computed among counties with non-zero Amish share. Holmes County (the outlier in Q4) is highlighted with a colored point and connecting line.

After detecting moderate positive spatial autocorrelation in LACV-ND incidence (Moran’s *I* = 0.35, *p* = 1.42e-9), we examined the local spatial structure using Local Indicators of Spatial Association (LISA). A statistically significant High-High cluster emerged in east-central Ohio (**Fig. S4**), indicating that counties with elevated LACV-ND incidence were spatially adjacent to other high-incidence counties. The Moran scatterplot (**Fig. S5**) confirmed this pattern, with several counties in the upper-right quadrant showing both high local incidence and high spatial lag values (*i.e.*, high average incidence among neighbors). This spatial structure motivated the use of spatial regression models to properly account for geographic dependencies in the data

We next modeled LACV-ND incidence by proportional Amish population using a spatial Durbin specification. Models with and without Holmes County’s inclusion showed that Amish population remained a significant predictor of LACV-ND incidence after adjusting for spatial structure, indicating it is not an artifact of spatial clustering (**Table 2**).

To assess whether Amish population effects persist beyond land use in a spatial framework, we next fit a full Spatial Durbin Model including Amish population share, deciduous forest cover, and pasture/hay cover as covariates, along with spatial lags of all three variables. This specification allows for both within-county effects and spillover effects from neighboring counties. We then decomposed effects into direct (within-county) and indirect (spillover) impacts for each covariate in the model. We found that the effect of Amish population share was significantly larger than any land-use variable (**Table 3**). Specifically, the total effect of Amish share (β_total_ = 8.2) substantially exceeded the total effects of deciduous forest (β_total_ = 0.3) and pasture/hay (β_total_ = -0.7), and confidence intervals for Amish share were entirely positive whereas land-use effects overlapped zero (**Fig. 3**). The indirect impact exceeded the direct impact, a result that implies that a county’s LACV-ND incidence is more strongly associated with its neighboring counties’ Amish populations than with its own, although the direct, or within-county, effect remained substantial (**Table 4**). The dominance of indirect over direct effects (indirect = 5.6 versus direct = 2.7) suggests that spatial spillovers play a major role in determining local LACV-ND incidence. This role of indirect effects potentially reflects mosquito dispersal, shared environmental conditions, or regional clustering of risk factors. Consistent with these results, both the Amish population share and LACV-ND case incidence are spatially clustered in central-eastern Ohio, with Holmes County as the area of highest concentration (**Fig. 4, S1, S3**). Counties high on both Amish share and incidence are concentrated in the east-central region surrounding Holmes County

**Figure 3:**
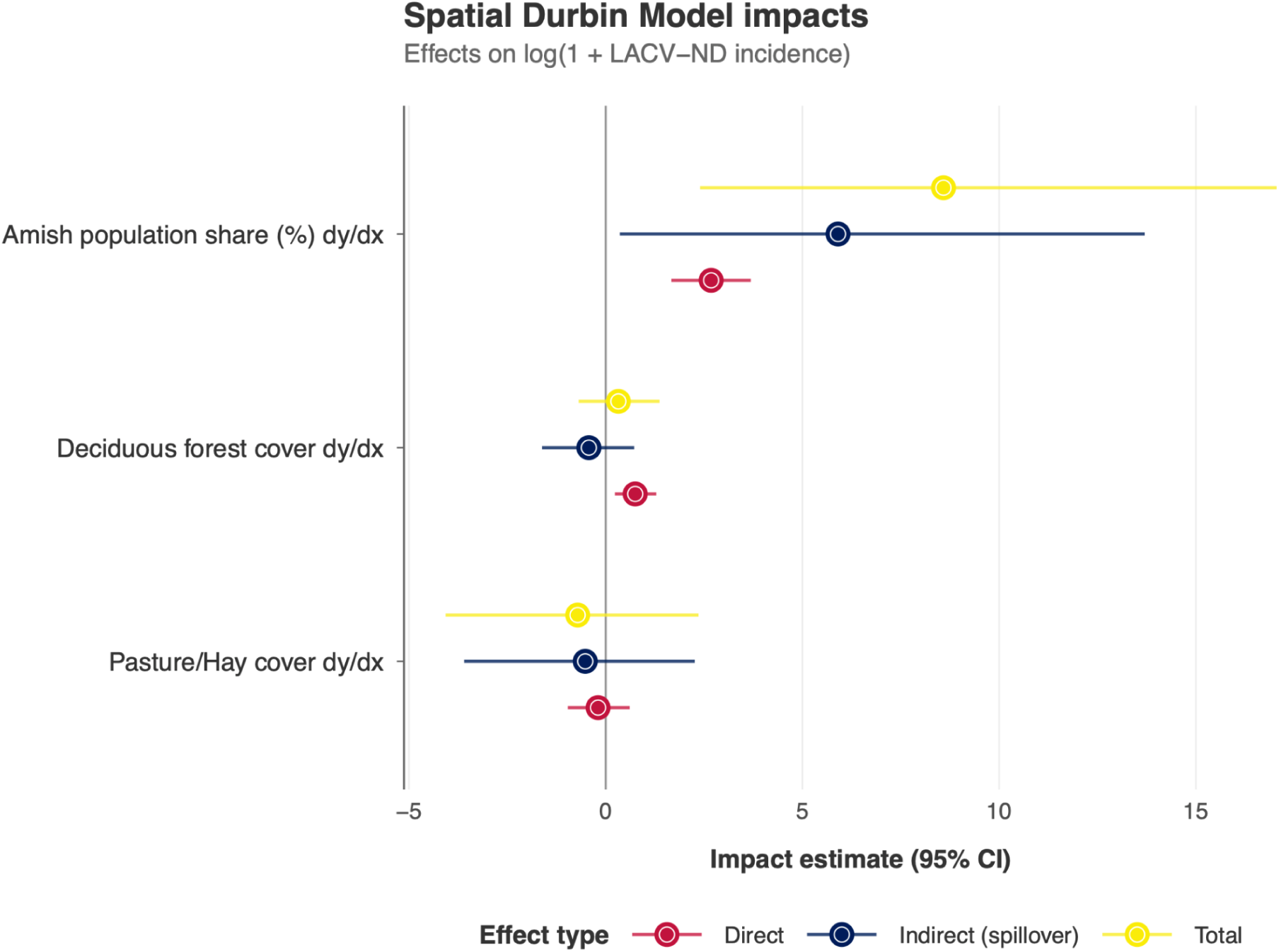
Spatial Durbin Model impact estimates for LACV-ND incidence. Forest plot displaying direct, indirect (spillover), and total effects of three covariates on log(1 + LACV-ND incidence). Point estimates are shown as colored circles with 95% confidence intervals as horizontal lines. Direct effects measure within-county impacts; indirect effects capture spatial spillovers to neighboring counties; total effects sum both components. Variables are ordered by magnitude of total effect, with Amish population share showing the largest positive impacts across all three effect types.

**Figure 4:**
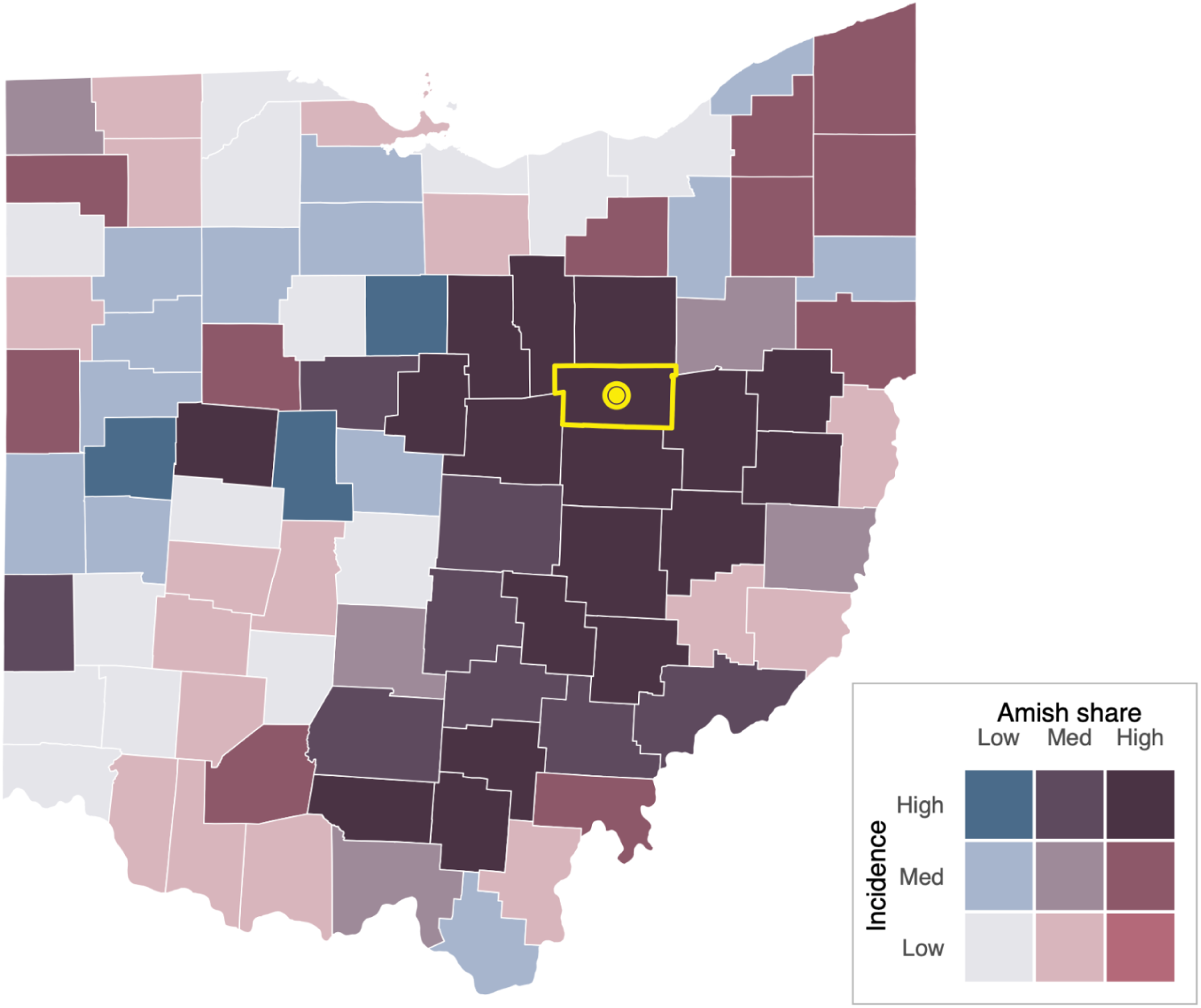
Bivariate choropleth of Amish population share and LACV-ND incidence in Ohio counties. Map displays the joint distribution of two variables using a 3×3 color scheme: Amish share increases from left to right (low to high) and incidence increases from bottom to top (low to high) in the legend matrix. Holmes County is outlined and marked with a central point.

**Table 3:**
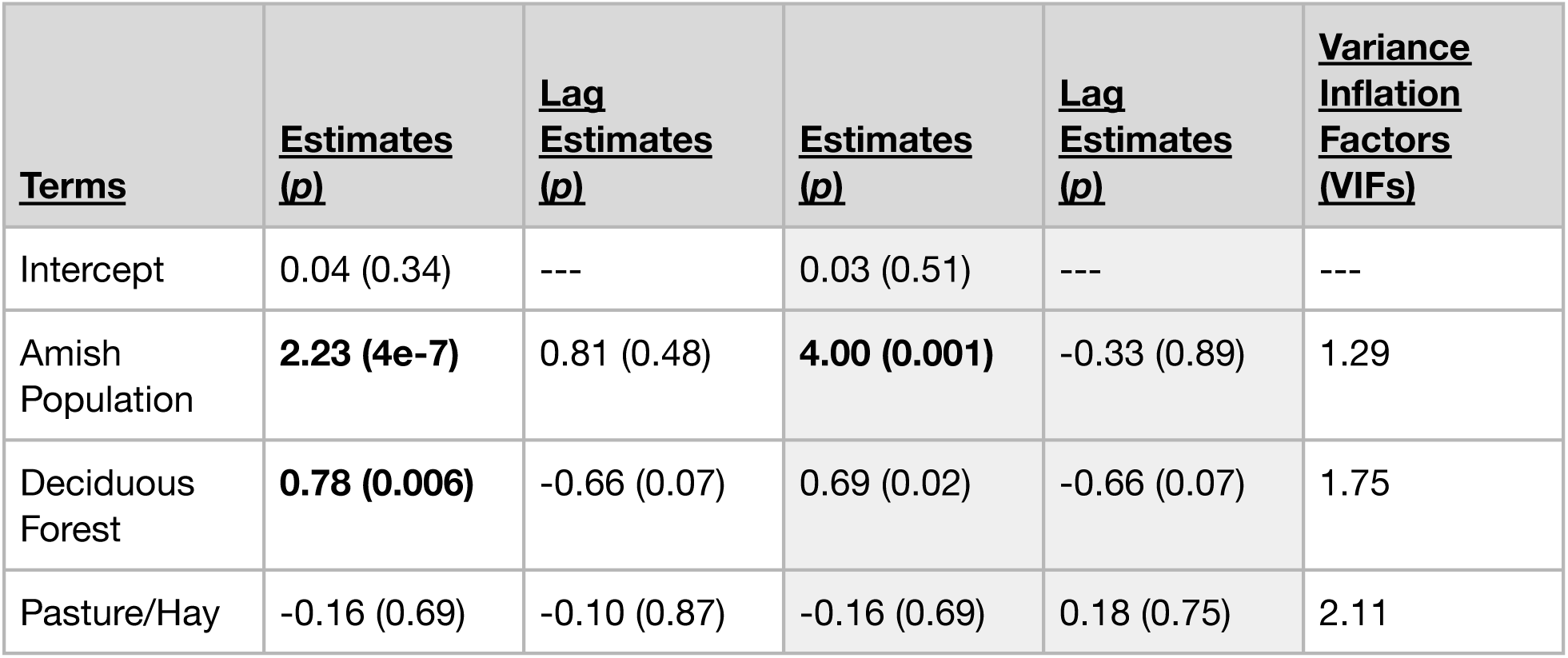
Summaries of spatial Durbin models. Bold text indicates the terms were significant at an alpha of 0.05. Shaded columns contain results of models that experimentally excluded Holmes County from the analysis. VIFs displayed here were calculated from an OLS model that included Holmes County.

**Table 4:** Impacts of terms in spatial Durbin models. Shaded columns contain results of models that experimentally excluded Holmes County from the analysis.

|  | **<u>Including Holmes Count</u>** |  |  | **<u>Excluding Holmes County</u>** |  |  |
| --- | --- | --- | --- | --- | --- | --- |
| **<u>Terms</u>** | **<u>Direct</u>** | **<u>Indirect</u>** | **<u>Total</u>** | **<u>Direct</u>** | **<u>Indirect</u>** | **<u>Total</u>** |
| Amish Population | 2.66 | 5.57 | 8.23 | 4.47 | 5.98 | 10.44 |
| Deciduous Forest | 0.75 | -0.42 | 0.33 | 0.65 | -0.54 | 0.11 |
| Pasture/Hay | -0.19 | -0.49 | -0.68 | -0.14 | 0.22 | 0.08 |

To evaluate model fit and assess whether spatial dependencies were adequately captured, we examined several diagnostic plots. Partial residual plots (**Fig. S6**) showed that the positive association between Amish share and incidence persisted even after accounting for spatial lags and land-use covariates, with Holmes County again appearing as an influential but consistent observation. The spatial distribution of model residuals showed no remaining spatial autocorrelation (Moran’s I = 0.007, Z = 0.286, p = 0.388; **Fig. S7**), confirming that the Spatial Durbin specification successfully accounted for geographic dependencies. The model achieved moderate in-sample fit (R² = 0.427, RMSE = 0.349 on log₁₊ scale; **Fig. S8**), with observed and predicted values clustering near the diagonal reference line.

Finally, to further demonstrate the importance of Amish population share in predicting LACV-ND incidence, we fit a reduced Spatial Durbin Model excluding the Amish covariate. This reduced model showed substantially degraded performance (R² = 0.191; **Fig. S9**), with particular underprediction of Holmes County’s incidence. When Amish share was excluded, the model severely underpredicted Holmes County incidence (observed = 2.95, predicted = 0.71), capturing only 24% of the actual value. The >50% reduction in explained variance when Amish share was excluded underscores its role as the primary driver of spatial variation in LACV-ND risk across Ohio counties.

We performed all statistical analysis in R ^38^, in which we used the following additional statistical packages: *car* ^39^*, sf* ^36,40^*, spatialreg* ^34^, and *spdep* ^35,36^. All code used in the analysis is available in a repository at https://github.com/bergeycm/Ohio-LACV-mapping-analysis.

## Discussion

La Crosse virus neuroinvasive disease (LACV-ND) is highly focal in Ohio despite near-ubiquitous vector presence, indicating the importance of other risk factors. In this analysis, we show that Amish population share is a robust predictor of LACV-ND incidence, more strongly associated with the disease case count than environmental variables summarizing land use. The indirect effect of Amish population share on LACV-ND incidence exceeded the direct effect, implying that the determinants proxied by Amish population share (e.g., household infrastructure, time outdoors, container ecology around homes, healthcare-seeking patterns) propagate across county borders. Our results suggest there is some anthropogenic ecological or socio-behavioral variation, proxied by Amish population share, that is amplifying or concentrating risk beyond what habitat suitability predicts.

The Old Order Amish are a Protestant denomination known for their distinct cultural practices ^41^. Many communities of Amish, known as settlements, are unelectrified to varying degrees and typically disconnected from external power grids, and they function without the use of automobiles ^42^. There is, however, notable variation in the use of modern technology among church districts, which are analogous to village-level groupings of multiple Amish families living in the same area ^41^. The Greater Holmes County settlement of Northeast Ohio has the distinction of being the second-most populous settlement in North America, closely following the Lancaster County area settlement in Pennsylvania ^43^.

Our findings suggest several plausible mechanisms for the association between LACV-ND incidence and Amish population share, and future work could test these hypotheses via co-developed, community-engaged evaluation. It is important to note that the community membership of the LACV-ND patients is unknown, but the association between Amish population share and LACV-ND cases still points to contextual, community-level conditions correlated with Amish settlement rather than the identity of individual patients. Potential mechanisms include *i.)* household infrastructure impacting exposure or creating larval habitats, and *ii.)* human behavior impacting exposure or diagnosis. First, household infrastructure may create conditions conducive to *Ae. triseriatus* reproduction and LACV transmission to people. LACV is transmitted by primarily day-biting mosquitoes (*Ae. triseriatus*, plus secondary vectors *Ae. albopictus* and *Ae. japonicus*), and exposure may be elevated where homes and schools have fewer window screens, less air-conditioning, and greater reliance on natural ventilation.

Infrastructure could also impact risk by introducing tree-hole analogs (e.g., rain barrels, stock tanks, buckets, cisterns, or tires), although this remains speculative. Second, human behavior may influence risk. Although farming remains important in many communities, many Amish households have diversified into trades and small businesses. Regardless, Amish children likely spend much more time outdoors compared to other Ohioan children ^42^. Finally, differences in care-seeking or pediatric LACV recognition may modify observed incidence. Some studies suggest lower use of conventional medical services among Amish ^44^, including in Holmes County, Ohio ^45^, which would tend to under-ascertain cases and making our observed positive association conservative if true. Clarifying these pathways will require community-based clinical and surveillance data.

Our statewide, multi-year dataset with explicit spatial modeling and impact decomposition is well-suited to quantify county-level associations. However, several factors limit our power to explain LACV-ND incidence fully. First, the analysis is constrained by coarse spatial resolution: disease incidence is aggregated to the county level and land-use metrics are similarly coarse; contemporaneous entomological data such as mosquito abundance or infection rates were unavailable. Second, we are reliant on reported cases of a nationally notifiable condition. Under-diagnosis and variable detection may bias observed incidence across counties. Third, unmeasured confounding variables likely co-vary with Amish population share. Such factors may include socioeconomic status, age structure, household size, healthcare utilization, and diagnostic capacity. Finally, results may be sensitive to model specification, including the choice of spatial weights and potential temporal mismatches between covariates and case years.

Our work points to several avenues for clarifying the observed association between Amish population share and LACV-ND, and for understanding how anthropogenic land use and the built environment shape transmission more generally. Foremost are within-county, high-resolution studies of hosts, vectors, and virus, particularly in Holmes County and neighbors. At the household- or parcel-scale, habitat audits could quantify tree holes and their analogs, ideally paired with microclimate logging. Fine-grained vector data would clarify local transmission ecology, including adult abundance and infection rates for *Ae. triseriatus* and secondary vectors, ovitrap indices, and bloodmeal identification to estimate the human blood index. In parallel, serologic surveys of humans and amplifying hosts (e.g., squirrels, chipmunks) could identify hotspots and allow estimation of force of infection. All determinants should be assessed using community-engaged research in collaboration with Amish and non-Amish participants to ensure feasibility and ultimately inform acceptable, effective interventions.

Like all vector-borne diseases, LACV-ND risk in Ohio reflects interactions between habitat and human ecology. Accounting for spatial dependence and removing an extreme outlier, Amish population share remains a strong predictor of LACV-ND. This suggests potential modifiable exposure pathways related to the built environment and human behavior. Focusing surveillance and control efforts at the interface of vector and human ecology offers a fruitful path to reduce LACV-ND in high burden areas such as Holmes County, Ohio.

## Data Availability

All code used in the analysis is available in a repository at https://github.com/bergeycm/Ohio-LACV-mapping-analysis.

## Acknowledgments

The authors would like to thank the Ohio Department of Health, especially their Zoonotic Disease Program. MEC thanks Caylee Heiremans and Peter Chaney for their continued support.

